# The benefit of a complete reference genome for cancer structural variant analysis

**DOI:** 10.1101/2024.03.15.24304369

**Authors:** Luis F Paulin, Jeremy Fan, Kieran O’Neill, Erin Pleasance, Vanessa L. Porter, Steven J.M Jones, Fritz J. Sedlazeck

## Abstract

The complexities of cancer genomes are becoming more easily interpreted due to advancements in sequencing technologies and improved bioinformatic analysis. Structural variants (SVs) represent an important subset of somatic events in tumors. While detection of SVs has been markedly improved by the development of long-read sequencing, somatic variant identification and annotation remains challenging.

We hypothesized that use of a completed human reference genome (CHM13-T2T) would improve somatic SV calling. Our findings in a tumour/normal matched benchmark sample and two patient samples show that the CHM13-T2T improves SV detection and prioritization accuracy compared to GRCh38, with a notable reduction in false positive calls. We also overcame the lack of annotation resources for CHM13-T2T by lifting over CHM13-T2T-aligned reads to the GRCh38 genome, therefore combining both improved alignment and advanced annotations.

In this process, we assessed the current SV benchmark set for COLO829/COLO829BL across four replicates sequenced at different centers with different long-read technologies. We discovered instability of this cell line across these replicates; 346 SVs (1.13%) were only discoverable in a single replicate. We identify 49 somatic SVs, which appear to be stable as they are consistently present across the four replicates. As such, we propose this consensus set as an updated benchmark for somatic SV calling and include both GRCh38 and CHM13-T2T coordinates in our benchmark. The benchmark is available at: 10.5281/zenodo.10819636 Our work demonstrates new approaches to optimize somatic SV prioritization in cancer with potential improvements in other genetic diseases.

## Introduction

Advancements in long-read sequencing technologies (LRS) have delivered unprecedented insights into cancer genomics (Aganezov et al. 2020; Akagi et al. 2023; Fujimoto et al. 2021; Choo et al. 2023; Thibodeau et al. 2020). Structural variants (SVs), defined as insertions, deletions and rearrangements larger than 50 base pairs (bp), represent an important subset of somatic driver events in tumours (Espejo Valle-Inclan et al. 2022; Aganezov et al. 2020). Examples include the amplification of oncogenes (eg. *ERBB2*) (Nattestad et al. 2018), generation of oncogenic fusion genes (e.g. *BCR-ABL* (Druker et al. 2001)), and silencing of tumour suppressor genes (e.g. *TP53* (Hernández Borrero and El-Deiry 2021)). Accurate detection of somatic SVs and characterization of their functional effects is thus critical for appropriate cancer diagnosis and selection of therapy. Short-read whole genome sequencing is routinely used for this purpose (Pleasance et al. 2020; Tsang et al. 2021), and can detect the majority of SVs (Choo et al. 2023). However, short-read sequencing is often unable to fully characterize the function of oncogenic SVs (Thibodeau et al. 2020; Pleasance et al. 2020). Even worse, short-read sequencing reports a high proportion of falsely identified SVs, which can be misleading (e.g. repeat expansions as translocations) (Thibodeau et al. 2020; Sedlazeck et al. 2018; Mahmoud et al. 2024, 2019). Cell lines (eg. SKBR3) have been analyzed to assess the complexity behind these oncogenic somatic changes. For example, Akagi et al. recently identified virus-mediated progression in head and neck cancer samples through the generation of unstable human papillomavirus (HPV) integrated molecular structures (Akagi et al. 2023). Further research is needed to assess the stability of SVs in cancer genomes, which can help direct priority when analyzing genomics data in heterogeneous cancer samples.

A key challenge in disease research is variant prioritization to identify potential causative variants of a condition. This is even more amplified in cancer patients since, depending on the type of cancer, the number of mutations are increased compared to normal tissue. Thus, researchers leverage normal controls (often blood) from patients to identify somatic variants that could potentially be driving the cancer (Mandelker and Ceyhan-Birsoy 2020). This process is efficient but also often complicated by multiple factors such as tumor purity, availability of non-cancerous tissue, gene annotation accuracy, and variant comparison/representation (English et al. 2022; Salzberg 2019; Yoshihara et al. 2013). In particular, variant prioritization is impacted by falsely identified variants in either tumor or normal samples. The cause of these falsely identified variants can be attributed to misinterpretation of mapped reads, coverage fluctuations, low quality mapping in tandem repeat regions, and unresolved regions in the reference genome. To improve these, multiple advancements have been proposed. The first complete reference genome CHM13-T2T recently became available, which showed improvements in variant calling (Nurk et al. 2022; Aganezov et al. 2022), and even corrected mis-represented medically relevant genes (Behera et al. 2023). The CHM13-T2T genome added approximately 200 mega base pairs (Mbp) of resolved sequence to the GRCh38 reference build, thus closing reference gaps and completing the centromere and telomere sequences (Nurk et al. 2022). These repetitive regions can play a key role in cancer progression as they include hundreds of protein coding genes that so far are not well understood (English et al. 2023). Perhaps more importantly, they can lead to the onset of genome instability and thus the formation of variants or inclusions of viruses (e.g. HPV) (Porter et al. 2023; Akagi et al. 2023). Annotating these newly assembled regions is challenging, but such annotations are necessary to understand the impact of mutations in these regions. While there are significant improvements in variant calling when SVs are detected against the CHM13-T2T reference (Aganezov et al. 2022), the benefits of using the complete genome reference have yet to be determined for somatic variants in a cancer context. Additionally, since CHM13-T2T is still being annotated, the annotated reference genomes GRCh37 and GRCh38 still hold value for ranking and characterizing SVs (Collins et al. 2020; Tanner et al. 2024). Using both a complete and annotated reference genome would allow researchers to identify novel oncogenic candidate variants across different cancer types.

Bioinformatics methodologies are continuously undergoing improvements to better utilize these novel reference genomes and enable improved detection of novel alleles, genes, and the variants and mutations that impact them (Smolka et al. 2024; Chen et al. 2024; Majidian et al. 2023). One such improvement was a novel method (Leviosam2) (Chen et al. 2024) that lifts over the read alignments between two reference genomes, thus gaining benefits of both and resulting in improved variant detection overall. This contrasts to previous methods that instead lifted over variant calls, which often lead to false positive variant calls, especially with larger rearrangements or duplications (Aganezov et al. 2020). We recently developed the SV caller Sniffles2 to identify SVs using long-read sequencing data (Smolka et al. 2024). While Sniffles2 has been applied in germline contexts such as neurological and mendelian disorders, its utility for cancer is still unproven, and somatic mutation detection often requires additional steps to reduce the false discovery rates. Furthermore, widely available benchmark samples are important instruments for benchmarking approaches (Zook et al. 2020; Olson et al. 2023; Majidian et al. 2023; Wagner et al. 2022). Over the past years several groups have proposed benchmarks for normal and cancer samples such as SKBR3 and COLO829 (Craig et al. 2016; Espejo Valle-Inclan et al. 2022). Nevertheless, these require constant vetting as reference genome changes and novel technologies change the downstream results that constitute the established benchmark.

In this work, we investigated novel advancements in SV calling and reference genome mapping to improve variant prioritization of somatic structural rearrangements. To accomplish this, we analyzed four replicates of the tumour/normal benchmark samples COLO829/COLO829BL at different laboratories to investigate the genome stability of these samples and also investigate how mapping to a complete reference genome (CHM13-T2T) influences the established benchmark. This highlighted a reduction of falsely identified SVs across all COLO829/COLO829BL samples when using the CHM13-T2T reference. This observation was also replicated in POG patient cancer samples where we observed a slight decrease in the number of somatic SV. Nevertheless, the lack of annotations on CHM13-T2T complicates variant prioritization. To overcome this, we propose a liftover approach of aligned reads that combines the benefits from both reference genome versions to deliver less falsely identified variants on GRCh38, further benefiting from the vastly available annotation resources. In addition to this work, we further investigated the genome instability of COLO829/COLO829BL and thus the risk of utilizing previous postulated benchmarks. We highlight multiple falsely reported and missed somatic SVs for this cell line on GRCh38. Furthermore, we propose a corrected, stable benchmark based on the four replicates of COLO829/COLO829BL sequenced across GRCh38 and CHM13-T2T. This should improve future cancer studies that use the CHM13-T2T reference genome for their analyses, which we found improves variant detection and prioritization overall.

## Results

### An updated COLO829 structural variation benchmark

We investigated the previously established COLO829/COLO829BL benchmark (Espejo Valle-Inclan et al. 2022), as recent reports highlighted discrepancies between the current benchmark and other sequencing data (Smolka et al. 2024; Shiraishi et al. 2023). We utilized four independent COLO829/COLO829BL samples from four sequencing centres profiled using the ONT and PacBio long read platforms, including the reads from the most recently established benchmark (Espejo Valle-Inclan et al. 2022) (**Supplementary Table 1**). **Figure 1A** shows the steps followed to obtain somatic SV calls that were used to investigate the aforementioned benchmark dataset (see Methods). Briefly, we aligned the four tumor/normal pairs to the GRCh38 reference genome, then identified SVs using Sniffles2’s population call/merge strategy (Smolka et al. 2024). We identified 19,684 SVs shared among all samples **(Figure 1B**), of which 45 were unique to the COLO829 cancer cell lines. Notably, the COLO829/COLO829BL samples by Valle-Inclan (labeled VAI) had the largest numbers of unique SVs (n_tumor_ =200, n_normal_=193) while the tumor-normal pair sequenced with the PacBio Revio platform (labeled PBR) has the lowest number (n_tumor_=38, n_normal_=65), which could be attributed to either differences in sequencing technology (Mahmoud et al. 2024) (older version of ONT MinION vs. PacBio Revio) or cell-line divergence. Interestingly, we observed that a small number of SVs were shared only by the tumor/normal pairs at individual sequencing centers (i.e detected in both COLO829 and COLO829BL but only in one of the four datasets). The lowest number of COLO829/COLO829BL shared SVs by sequencing center was 15 from Canada’s Michael Smith Genome Sciences Centre (labeled as GSC) and the highest was 449 from the Valle-Inclan (ONT 271, PBR 115). Finally, there were 8,659 SVs that were shared between two to seven samples (tumor or normal) from different platforms and sequencing centers, which were omitted from subsequent analysis (**Supplementary Table 2**). We filtered for SVs that were present in the four cancer samples with a variant allele frequency (VAF) >= 10% and not present in any of the normal samples. Subsequently, the remaining somatic 44 SVs were manually reviewed in IGV. We denoted this dataset as COLO829-GRCh38 (**Supplementary Table 3**).

**Figure 1.**
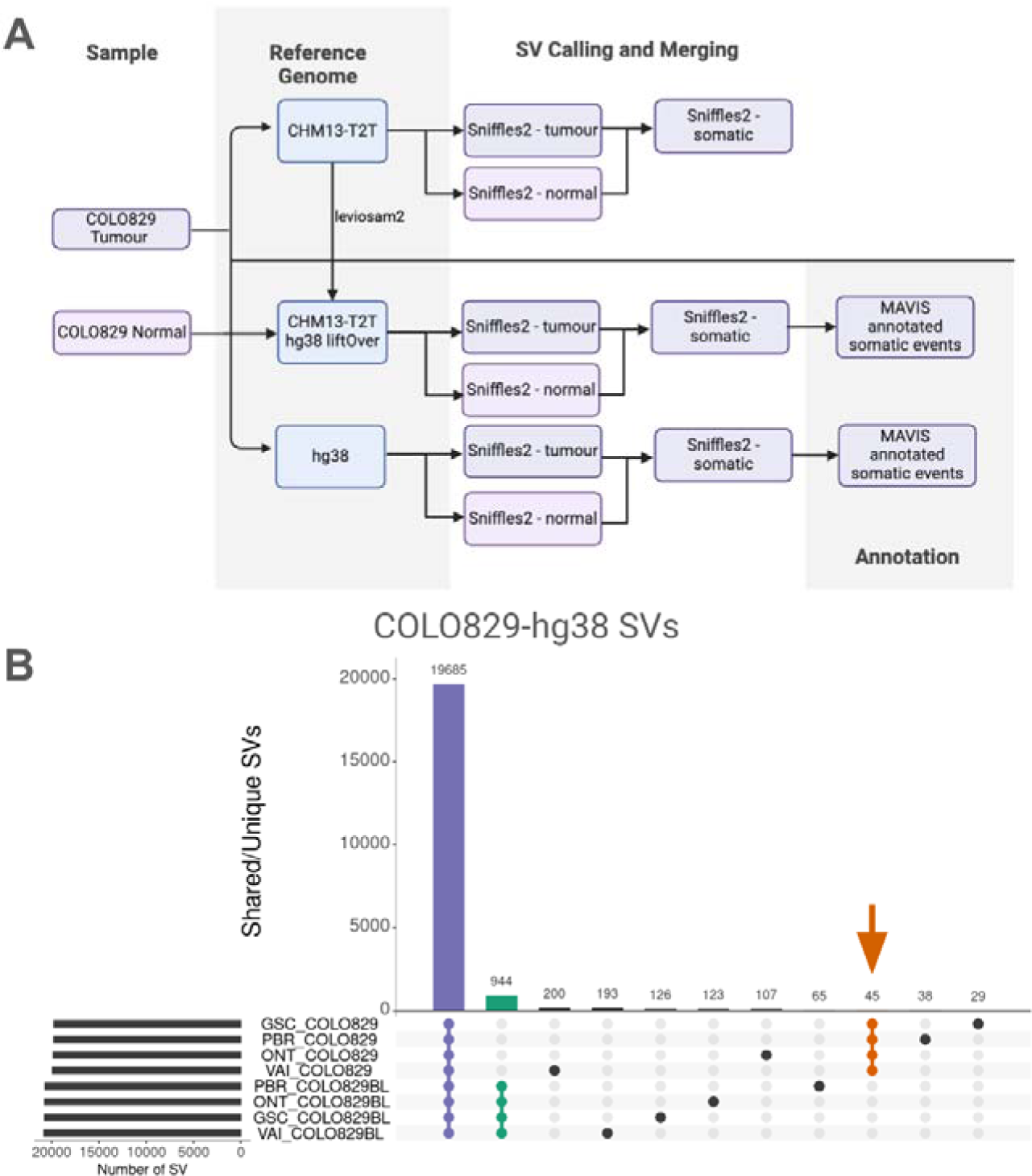
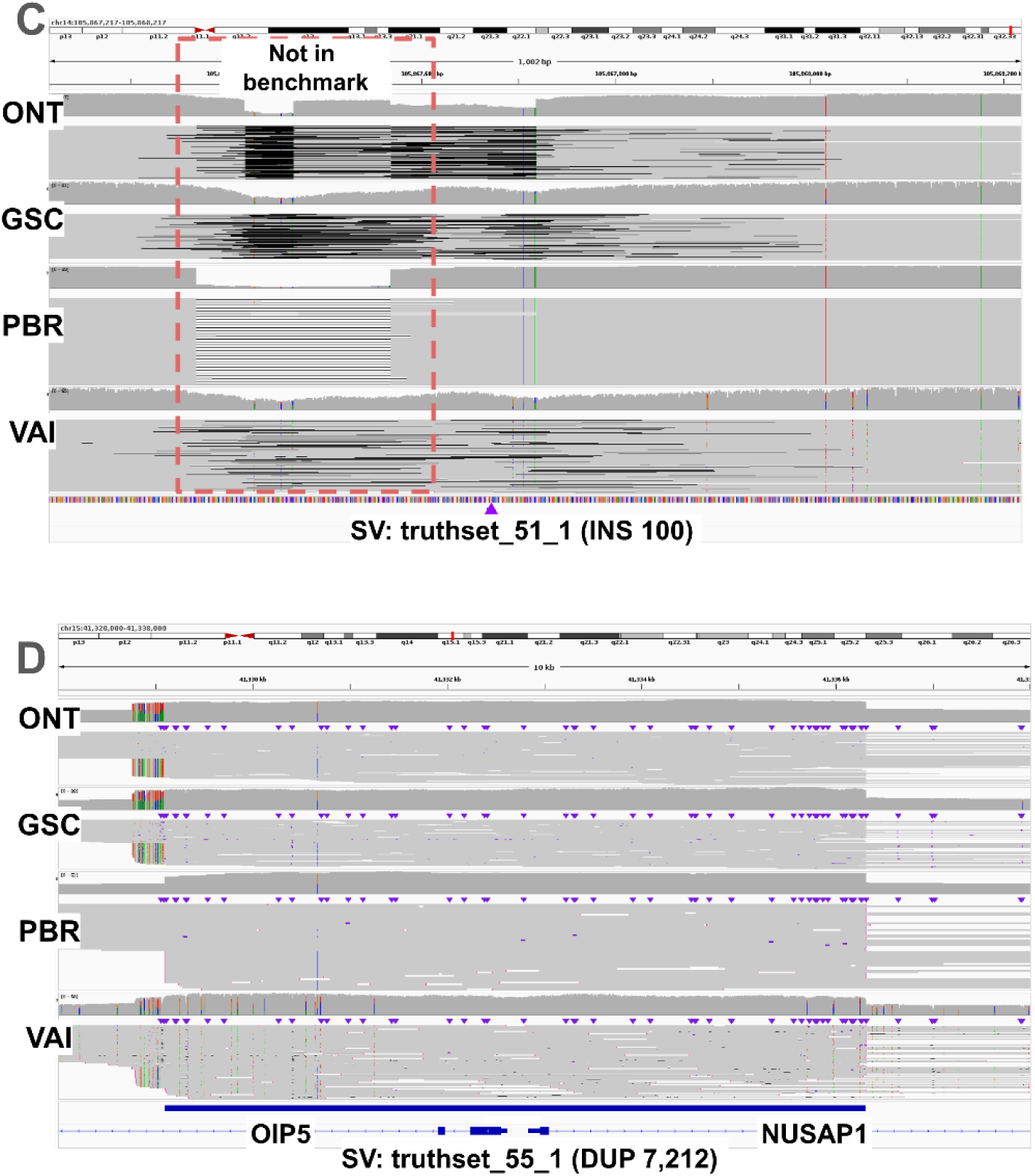
**A)** Schematic overview of our variant calling and comparison methods using CHM13-T2T v GRCh38. **B)** Upset plot of the shared and unique SVs of the COLO829-GRCh38 dataset across four COLO829 tumor/normal pairs. Violet events shared by all samples; orange, somatic events shared by tumour samples only; green, germline events shared by blood normal samples only. **C)** IGV screenshot of an SV cataloged as missing (truthset_51_1, large purple triangle) according to the benchmark by Valle-Inclan et al. We did not detect any reads supporting it. Moreover, we detected a DEL in all four sample which is missing from the benchmark. **D)** IGV screenshot of a DUP that was assigned two distinct SV types (DUP and INS), and thus cataloged as missing according to the benchmark by Valle-Inclan et al. Manual inspection in IGV showed the SV.

We compared these 44 SVs called in data from all four centers to the previously established SV benchmark for COLO829 (62 SVs, see Methods) by Valle-Inclan et al. (Espejo Valle-Inclan et al. 2022). **Table 1** shows the benchmark summary. In comparison to established benchmark SVs, we categorized 38/44 SVs as true positives (TP), six as false positives (FP), and 24 as false negatives (FN) (**Supplementary Table 4**). We proceeded with an in-depth analysis of the FP and FN calls to understand how the previous benchmark compared to the full set of data from four centres. A manual inspection of FP events revealed that 5/6 were falsely classified as FPs as they showed clear evidence in samples from multiple centres (**Supplementary Table 5, Supplementary Figure 1**). For example, the variant INS.C2M6, a heterozygous 69 bases insertion, which involved *PMS2*, was clearly detected in all four cancer samples (**Supplementary Figure 1B**) but was not reported in the previous benchmark (Espejo Valle-Inclan et al. 2022). For the 24 SVs classified as FN, we performed a genotyping experiment (Chander et al. 2019) (also known as force-calling, see Methods, **Supplementary Table 6**) in order to assess if there was any signal in the data to support such SVs. Here, five SVs that were initially missed by our analysis had strong read evidence in all four COLO829 replicates (**Figure 1D, Supplementary Figure 2**), one FN SV only was detected in a control sample with one read thus was discarded by our filter that removed SVs with any evidence in the normal samples (**Supplementary Figure 3**). In addition, ten FN SVs had variant allele frequencies (VAF) below our threshold of 10% and therefore were not included (VAF 1.4-6.2%, **Supplementary Figure 4**), additionally, four FN SVs could only be detected in 1-3 COLO829 replicates, and thus were filtered out (**Supplementary Figure 5**), and finally, four SVs had no read support for the SVs across all four cancer replicates (**Figure 1C**, **Supplementary Figure 5C**). Interestingly, we missed a well defined deletion present in all four COLO829 replicates (benchmark SV truthset_56_1). Further investigations revealed that a filter applied during SV calling prevented the detection of that particular SV (**Supplementary Figure 2C**). This filter is applied to remove potential erroneous SVs based on the minimum coverage (COV_MIN) needed to detect an SV. In total, 43 somatic SVs detected in SV calling in COLO829-GRCh38, excluding one false positive, and 5 somatic SVs called from force-called genotyping and manual review had strong read evidence in datasets from all four centres.

**Table 1.** Benchmark results for the three COLO829 datasets based on the reference genome. For each dataset we present the initial evaluation (labeled Initial) and the in-depth analysis/evaluation that includes genotyping, and manual inspection in IGV.

| Datset | Version | P | TP | FP | FN | Precision | Recall |
| --- | --- | --- | --- | --- | --- | --- | --- |
| COLO829-38 | Initial | 62 | 38 | 6 | 24 | 86.36% | 61.29% |
|  | Genotyped/IGV | 49 | 44 | 1 | 5 | 97.78% | 89.80% |
| COLO829-T2T | Initial | 62 | 38 | 5 | n/a | 88.37% | n/a |
|  | Genotyped/IGV | n/a | 43 | 0 | n/a | 100.00% | n/a |
| COLO829-lifted | Initial | 62 | 36 | 5 | 26 | 87.80% | 58.06% |
|  | Genotyped/IGV | 49 | 45 | 0 | 4 | 100.00% | 91.84% |

### Leveraging a complete genome reference for cancer structural variation benchmarking

The recent introduction of CHM13-T2T showed improved variant calling (Aganezov et al. 2022; Nurk et al. 2022) and corrections of a few medically important genes (Behera et al. 2023). Thus, we examined the impact of using CHM13-T2T for structural variant calling in cancer research. Using the CHM13-T2T, we observed a high number of SVs in the telomeric and centromeric regions of the chromosomes, the detection of which was aided by a complete reference (**Supplementary Figure 6**). **Supplementary Table 8** shows the comparison of SVs detected in centromeric and telomeric regions between using the CHM13-T2T reference (labeled as COLO829-T2T) and COLO829-GRCh38. From COLO829-T2T, 18 chromosome have higher number of SVs in centromeric and telomeric regions (average 16.55% increase), while six chromosomes have higher number of SVs in centromeric and telomeric regions in COLO829-GRCh38 (average 1.76% increase). Next, when somatic SVs are identified in the COLO829-T2T dataset (**Supplementary Table 7**), using the same methodology as described for the benchmark SV set, we do not observe an enrichment in such regions (red dots in **Supplementary Figure 6**), suggesting these events are mostly germline variation. We observed a deviation in the ratio of insertions (INS) and deletions (DEL) with an INS:DEL ratio of 1:1.4 in CHM13-T2T and 1:0.87 in GRCh38. The switched INS:DEL ratio has been described before and is thought to be caused by too small tandem repeats reported in GRCh38 (Aganezov et al. 2022). Furthermore, we observed a substantial decrease in the number of interchromosomal events (BND) from 225 in GRCh38 to 83 in CHM13-T2T for COLO829. As interchromosomal events are not expected to be found in normal samples, this reduction largely reflects a decrease in noise in variant calling.

To enable a comparison between the GRCh38 and CHM13-T2T based SV calls, we linked SV calls of the same SV type and chromosome using the read names supporting each SV. Since there is no COLO829 SV benchmark with CHM13-T2T coordinates, we used the read names to determine which SVs correspond to the benchmark based on the previous genotyping. Figure 2A shows the number of SVs shared across the sample replicates. The total number of germline SVs was slightly smaller in CHM13 compared to GRCh38 (19,314 and 19,685 respectively), while the number of events considered somatic and shared between tumour samples only was slightly larger (50 and 45 SV, respectively). Unlike the GRCh38 calls, here the samples from the Oxford Nanopore Open Data project (labeled as ONT) had the largest numbers of unique SVs (517, 242 for COLO829/COLO829BL) while the COLO829/COLO829BL sequenced by the GSC has the lowest number (33, 38 for COLO829/COLO829BL). Finally, there were 17,395 SVs that were shared between two to seven samples (tumor or normal) from different platforms and sequencing centers (**Supplementary Table 9**). These SVs were not included in the analysis. We manually reviewed the 50 SVs detected in tumour samples only from all four centres and identified 43 somatic SVs. Out of which 38 were initially classified as TP and five as FP relative to the comparison of the COLO829-GRCh38 somatic SV benchmark by Valle-Inclan et al (see Methods). We identified two interesting cases where the somatic SV is present in both GRCh38 and CHM13, but is called as being a different size due to differences in the references. SV DEL.492M6 differed in size by ∼6kb (Figure 2C) to the corresponding SV in GRCh38 SV truthset_28_1 from the benchmark by Valle-Inclan et al), overlaps with the glutamate metabotropic receptor *GRM8* and SV DEL.508M6 (corresponding to truthset_30_1 in the benchmark) differed in size by 126kb which in the case of CHM13-T2T SV is not hitting any genetic element but the GRCh38 does overlap with an Olfactory Receptor *OR2A1* and its antisense RNA *OR2A1-AS1*. This difference is a consequence of the size difference between both SVs and not given by differences in the annotation. Next, from the five somatic SVs initially classified as FP according to Valle-Inclan et al, SV DEL.1D9MB is uniquely detectable using the CHM13-T2T reference genome as it is positioned in the centromere of chromosome 12 (Figure 2B). Manual inspection of the remaining four FP showed that they actually represent TPs, as we detected supporting reads for all four sequenced COLO829 samples (**Supplementary Table 10, Figure 2D, Supplementary Figure 7**). For example, Figure 2D shows INS.570MD, a 97 bp insertion that can be clearly observed in all four cancer samples. Next, we used the UCSC genome browser (Kent et al. 2002) to annotate the somatic SVs for COLO829-T2T. Here, we observed similar results to the annotation of somatic SVs in GRCh38 coordinates (20/31, 64.5% shared genes), with the addition of nine characterized genetic elements and the centromere (**Supplementary Table 7B**).

**Figure 2.**
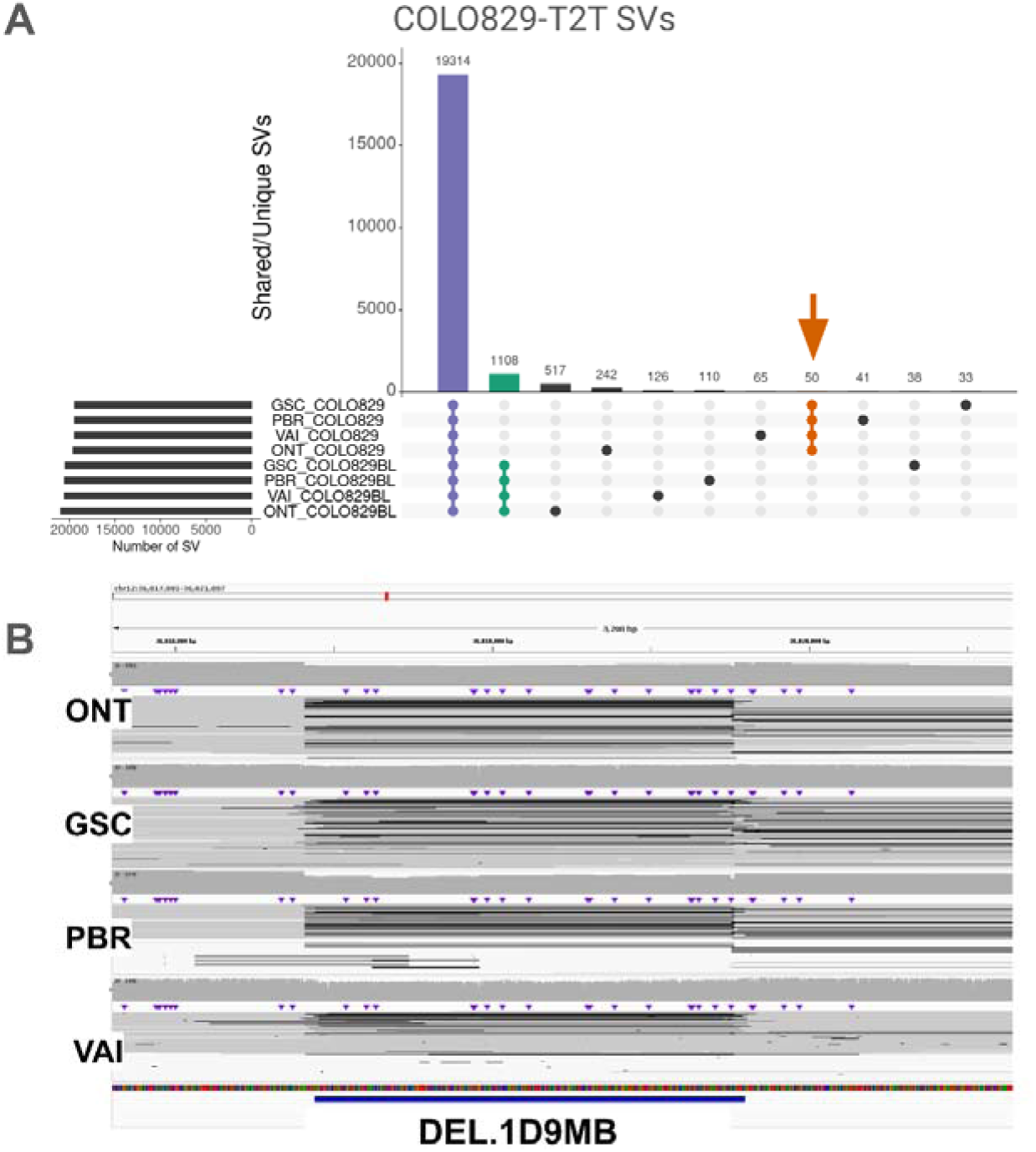
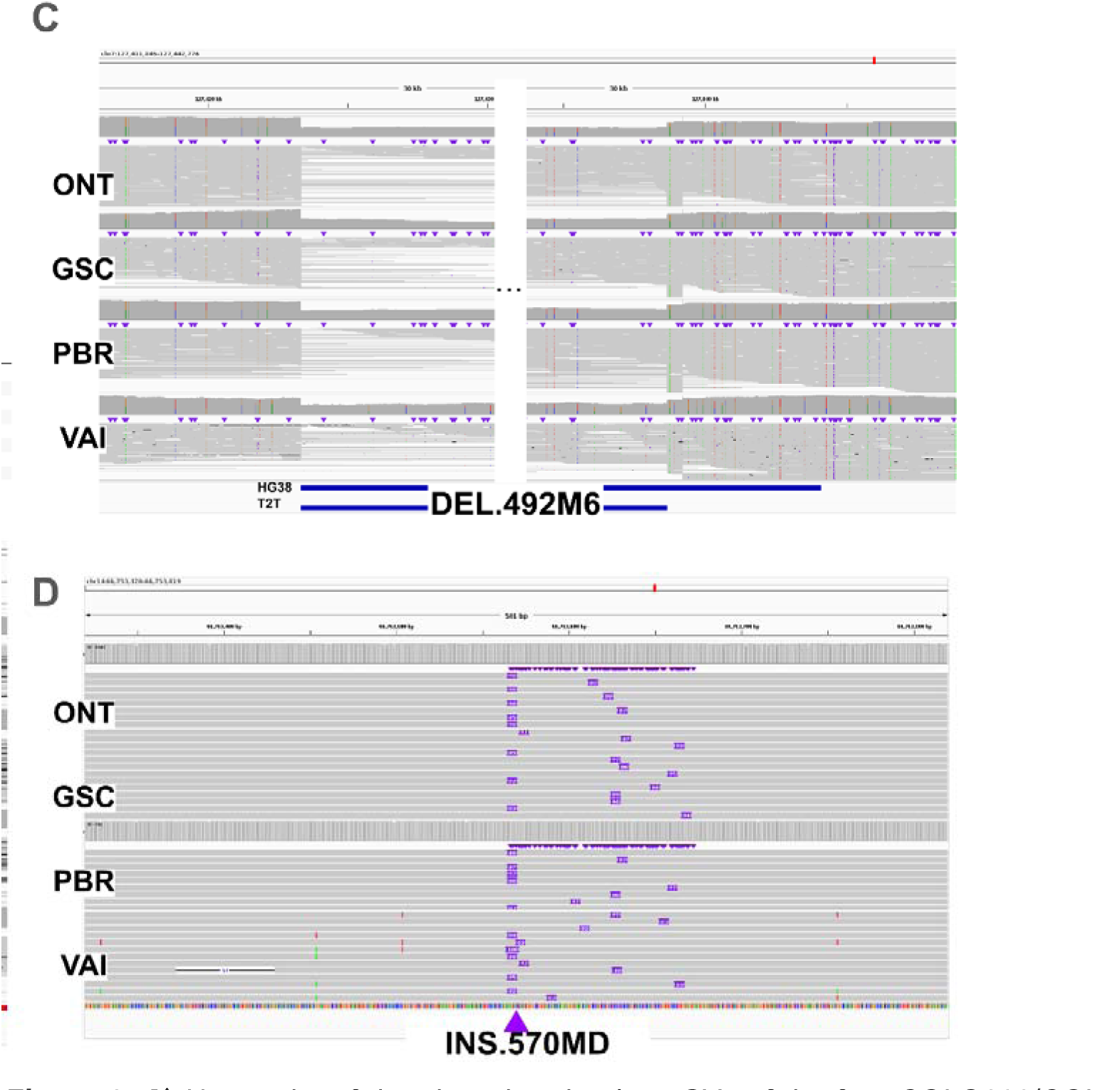
**A)** Upset plot of the shared and unique SVs of the four COLO829/COLO829BL tumor/normal pairs aligned to the CHM13-T2T reference genome. **B)** IGV screenshot of an SV that was reported with different sizes in CHM13-T2T and GRCh38. **B)** IGV screenshot of an SV detected in the centromere of chromosome 12 on CHM13-T2T. **D)** IGV screenshot of an SV cataloged as FP according to the benchmark by Valle-Inclan et al, which we are able to detect in all four cancer samples.

Although SV detection is improved (i.e. no FP SVs detected), using CHM13-T2T limits variant prioritization and interpretation as population frequencies are not available on this reference, and GRCh38 has more informative annotation databases (Collins et al. 2020; Nicholas et al. 2022; Chowdhury et al. 2022). Thus, we used recent advances in liftover of alignments to take advantage of both the improved mapping using a CHM13-T2T reference (Chen et al. 2024) and the years of annotation and curation of the GRCh38 reference genome. Briefly, we took the CHM13-T2T read alignments and used LevioSAM2 (Chen et al. 2024) (v0.4.1) to liftover the alignments from CHM13-T2T to GRCh38 coordinates. We denoted this dataset as “COLO829-lifted” (**Supplementary Table 11**). Figure 3A shows an upset plot with the number of SVs shared among all samples, those unique to the cancer cell line COLO829 (marked with an arrow), the blood control COLO829BL and the unique SV per sample. We compared the COLO829-lifted calls to the benchmark by Valle-Inclan et al, alongside the dataset COLO829-GRCh38 that was produced earlier. **Table 1** shows how leveraging the CHM13-T2T genome improves SV calling and decreases both the number of SV classified as FP and FN when compared to the native GRCh38 alignment, after genotyping and manual review (**Supplementary Table 12**). For the case of the SVs labeled as FP in comparison to the benchmark by Valle-Inclan et al, we were able to identify all the SVs as TP with high confidence in all the cancer samples (**Supplementary Table 13, Supplementary figure 8**). One interesting example is DEL.6A4M6, which is a deletion that was also called in COLO829-GRCh38 but with a greater length (46 bases in COLO829-lifted vs. 138 bases in COLO829-GRCh38); however, manual inspection in IGV shows the 46 base DEL only demonstrating the more accurate result obtained when incorporating T2T alignment. Next, when analysing the FN we identified two SVs that were also missed in the COLO829-GRCh38 (benchmark SVs: truthset_12_1 and truthset_56_1, **Supplementary Figure 9**). In both cases we applied filters during SV calling that removed them from the final call set (COV_CHANGE for the case of SV:truthset_12_1 and COV_MIN for SV:truthset_56_1). Then, two SVs labeled as FN were detected as INS in our callset and as DUP in the benchmark. These two types are often hard to distinguish as a duplication is an insertion of the same sequence next to itself (Mahmoud et al. 2019). The rest of the FN are very similar the COLO829-GRCh38 results (**Supplementary Figure 9-12**) with the difference of truthset_23_1 being missed in the GRCh38 dataset but not in the liftover and truthset_38_1 being classified as low-allele frequency (VAF < 10%) in the GRCh38 and present in the liftover.

**Figure 3.**
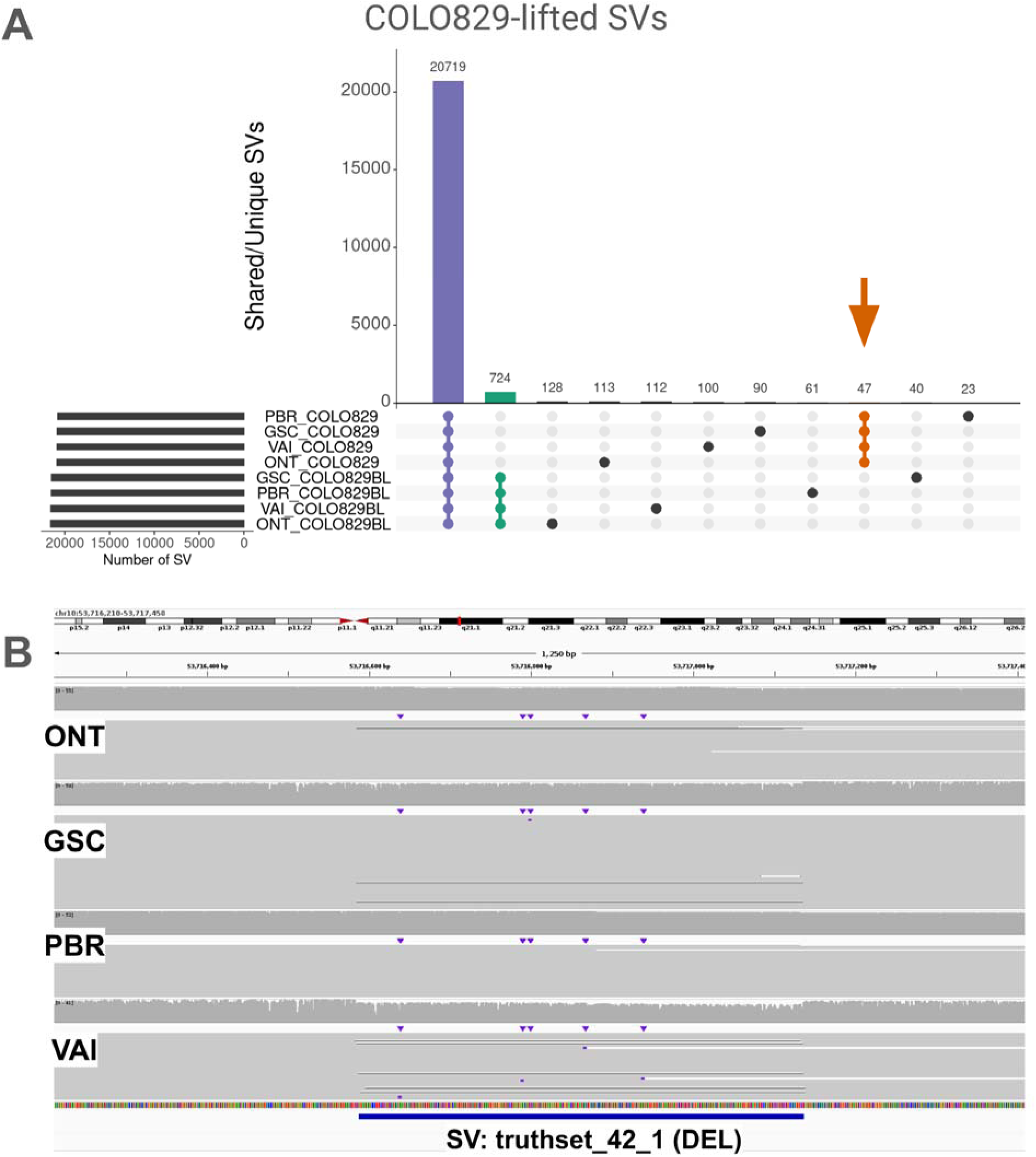
**A)** Upset plot of the shared and unique SVs of the four COLO829 tumor/normal pairs aligned to the CHM13-T2T reference genome and lifted over to GRCh38 (**Supplementary Table 14**). **B)** IGV screenshot of an SV that was labeled as FN according to the benchmark by Valle-Inclan et al. Our analysis calls removed thee SVs due to lack of evidence in all samples (**Supplementary Table 13**)

Overall, the approach of incorporating alignment to CHM13-T2T followed by liftover analysis eliminated all FP somatic SV calls for COLO829. Such a reduction in FP has important impacts particularly when analysing patient samples, where a reduction in FP can have implications for interpretation and necessary follow-up analysis. The SV liftover approach allows for combining the improved accuracy gained by incorporating CHM13-T2T alignments (precision in COLO829-GRCh38 97.78% vs COLO829-T2T/COLO829-lifted 100.00%) with the biological and clinical annotations available on GRCh38. Additionally, we observed a drastic reduction in BND (225 and 130 respectively for COLO829-GRCh38 and COLO829-lifted, 83 in COLO829-T2T), both in tumor and normal samples, indicating improvements in germline SV calling which has implications for variant prioritization in disease research. We used MAVIS (Reisle et al. 2019) to annotate our proposed somatic SVs for the COLO829 cell-line in GRCh38 space. From the 49 somatic SVs detected in COLO829-lifted (45 TP and 4 FN), 35 SVs overlap with 26 known genes, of which 25 are related to cancer. Some examples are *FHIT* (near a fragile site), tumor suppressors ITIH5, *MAGI2, PTEN, WWOX,* and cell-proliferation control gene *TMX3* (Bellon et al. 2021; Cao et al. 2020; Yehia et al. 2023; Husanie et al. 2022; Zhang et al. 2019).

Since these 49 somatic SVs could be detected across all 4 replicates of COLO829, we suggest using this call set as a refined benchmark for this important tumor/normal control cell line (**Supplementary Table 15**). The use of data from multiple centres and independent cell line passages is particularly important as we demonstrate differences between samples that are likely to in part represent instability and evolution of this cell line at the SV level in addition to the previously described instability at the SNV level (Craig et al. 2016). Thus, accurate benchmarks that incorporate multiple replicates are necessary to reduce possible sources of error introduced by a single sample.

### Utilizing a complete genome reference for cancer SV detection

Our results show the benefit of using the CHM13-T2T reference for SV analysis in cancer. Furthermore, we introduced and assessed a liftover approach that leverages the benefits of both reference genomes to improve the detection and annotation of somatic SVs in COLO829/COLO829BL. Additionally, variant prioritization can be challenging as most SVs are not cancer drivers, but instead passenger mutations picked up in the process of cancer evolution. Cancer samples have diversity in tumour content (fraction of sequenced DNA derived from tumour cells compared to normal cells) which can result in somatic SVs with low VAF. Because of the overall complexity of SV analysis in cancer, a reduction in false positive calls is greatly beneficial. Given these results, we next investigated the use of the CHM13-T2T reference genome for the analysis of two different cancer patient samples. In this pilot experiment, we used two samples with high whole genome coverage (52x and 54x) and matching normal controls (blood and skin) (**Supplementary Table 1**) (O’Neill et al. 2024). When using CHM13-T2T alignment followed by GRCh38-liftover analysis, we observed fewer somatic SVs when compared to GRCh38 (**Supplementary Table 17A-D**).

For POG044, we observed 193 SVs in the liftover analysis compared to 201 using the GRCh38 reference (**Supplementary Table 17A and 17B**), with a similar INS to DEL ratio (1.22:1 in the liftover, 1.23:1 in GRCh38). We observed a reduction in the number of INS, DEL and BND (96, 79, 13, to 100, 81, 16, respectively) and no difference in DUP or INV. For sample POG1022, we detected an increase in the number of DEL in both the liftover and the GRCh38 analysis (**Supplementary Table 17C and 17D**). Also, the INS to DEL ratio was skewed towards DEL (0.81:1 and 0.77:1 for liftover and the GRCh38 respectively). In a more detailed inspection we detected a high number of DEL in pericentromeric regions of chromosomes 4, 10 and 13 (eight, 36 and 14, respectively). Also, we detected no change in the number of BND, and a decrease in the rest of the SV types.

We annotated SVs from the liftover analysis (in GRCh38) using MAVIS, but overall did not observe any SV that would be predicted to be causative in tumour formation. MAVIS predicted a non-synonymous SV on chr1 in the sample POG044 (**Supplementary Table 18**). This SV affects the neuroblastoma breakpoint family gene *NBPF20* which has been associated with several types of cancer. This region was affected by several deletion events, moreover most of them show low mapping quality and thus were not used during SV calling, specially in the control sample (**Supplementary Figure 13**).

For sample POG1022, three non-synonymous coding events were categorized by MAVIS (**Supplementary Table 19**). The first one, a 24.2Kb inversion in chromosome 2 which affects the immunoglobulin kappa variable gene *IGKV3-11* (**Supplementary Figure 14**) which expression has been observed in myelomas (https://www.proteinatlas.org/ENSG00000241351-IGKV3-11/pathology. The second, a 8.9Kb deletion also in chromosome 2 which affects the ankyrin repeat domain gn *ANKRD36* (**Supplementary Figure 15**) which has been used as a biomarker of disease progression in Leukemia (Iqbal et al. 2021). Upon further inspection, we detected a larger deletion in the same region (12kb) with some reads supporting the SV in the control. The third non-synonymous coding event was a 609kb deletion in chromosome 20 which was interestingly only detectable in the lift over analysis and not in the GRCh38 alignment. Visual inspection of the region in both the liftover analysis and GRCH38 alignment shows that the latter has a weaker signal, although it is present. This SV affects the prostaglandin synthase *PTGIS* (Figure 4) which has been linked with various cancers like prostate cancer (Qiao et al. 2023), colorectal cancer (Ding et al. 2023) and cancer-free trichothiodystrophy (Lombardi et al. 2021).

**Figure 4.**
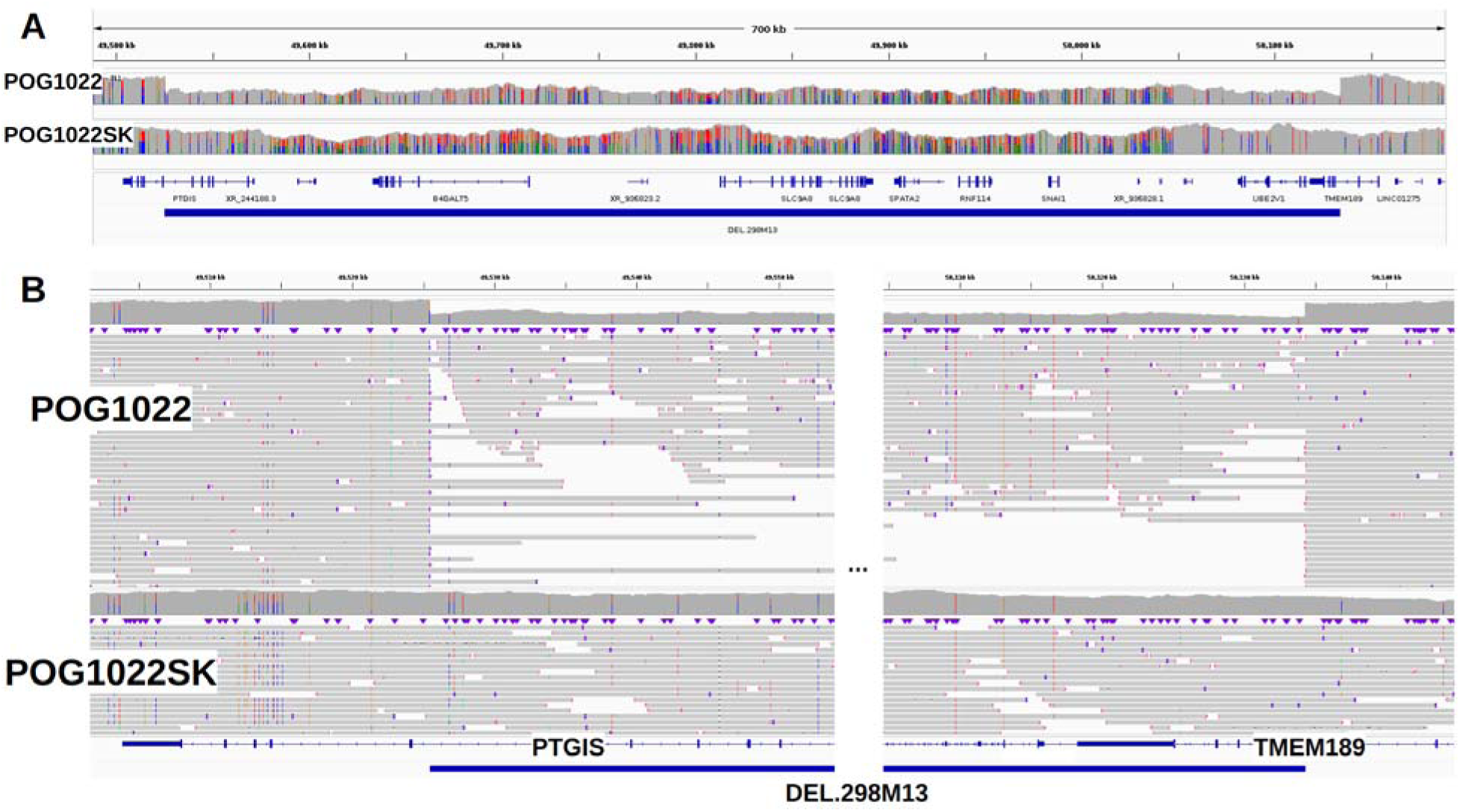
**A)** Coverage pattern for a 609kb somatic deletion (DEL.298M13) detected in POG1022 that affects PTGIS. (**Supplementary Table 18**). **B)** IGV screenshot that zooms into the breakpoints of the somatic deletion DEL.298M13.

Closer inspection of the 140 somatic SVs from POG044 that were annotated by MAVIS (44 were filtered or collapsed) showed that 96 (68.6%) were not somatic, meaning we could detect reads supporting the SV in the control samples. From those 30 were caused by SV of the same type that were collapsed in the tumor sample but not in the control causing a difference in size and thus two different SV (**Supplementary Figure 16A**), 23 were in low complexity regions where we could observe multiple SVs of the same type (**Supplementary Figure 16B**) and 18 have read support in the control with mapping quality lower than out threshold (MQ≥20) and thus were not used during SV calling (**Supplementary Figure 16C**). For sample POG1022, MAVIS annotated 184 somatic SVs (100 were filtered or collapsed) from which 129 (70.1%) had reads supporting the SV in the control sample. From those, 64 occurred in low complexity regions (**Supplementary Figure 17A**) and 46 were caused by collapsed SV of the same type (difference in size **Supplementary Figure 17B**).

Next, we investigated the SVs detected when using the CHM13-T2T. While we found CHM13-T2T somatic events in both POG044 (n = 407) and POG1022 (n = 433) (**Supplementary Table 20**), upon manual review, we could match 55 to GRCh38 in POG044 and 83 in POG1022 based on the read names. Moreover, even when we observed an increase in the number of INS and DEL using CHM13-T2T, in both cases we observed a reduction in the number of BNDs, while the rest of the SV types were comparable. Similar to our previous analysis with the COLO829 cell-line, we observed the INS:DEL ratio skewed towards deletions.

A closer inspection of the somatic events in these two samples from the POG cohort, we observed that most were not somatic, but lay in low complexity regions (i.e centromere/telomere but not exclusively). We detected reads with support of the SVs in the control samples (POG044BL and POG1022SK respectively) but with mapping quality below or established threshold, such that the combined with the mis-alignments, they caused INS, and mostly DEL SVs that were labeled as somatic. Thus, new advances in algorithms to overcome these previously inaccessible regions are needed. Furthermore, when removing the SV that were not in the aforementioned repetitive regions (centromere, telomere), we detected a lower number of SV than using the GRCh38 (Figure 5, **Supplement figure 18-19**)

**Figure 5.**
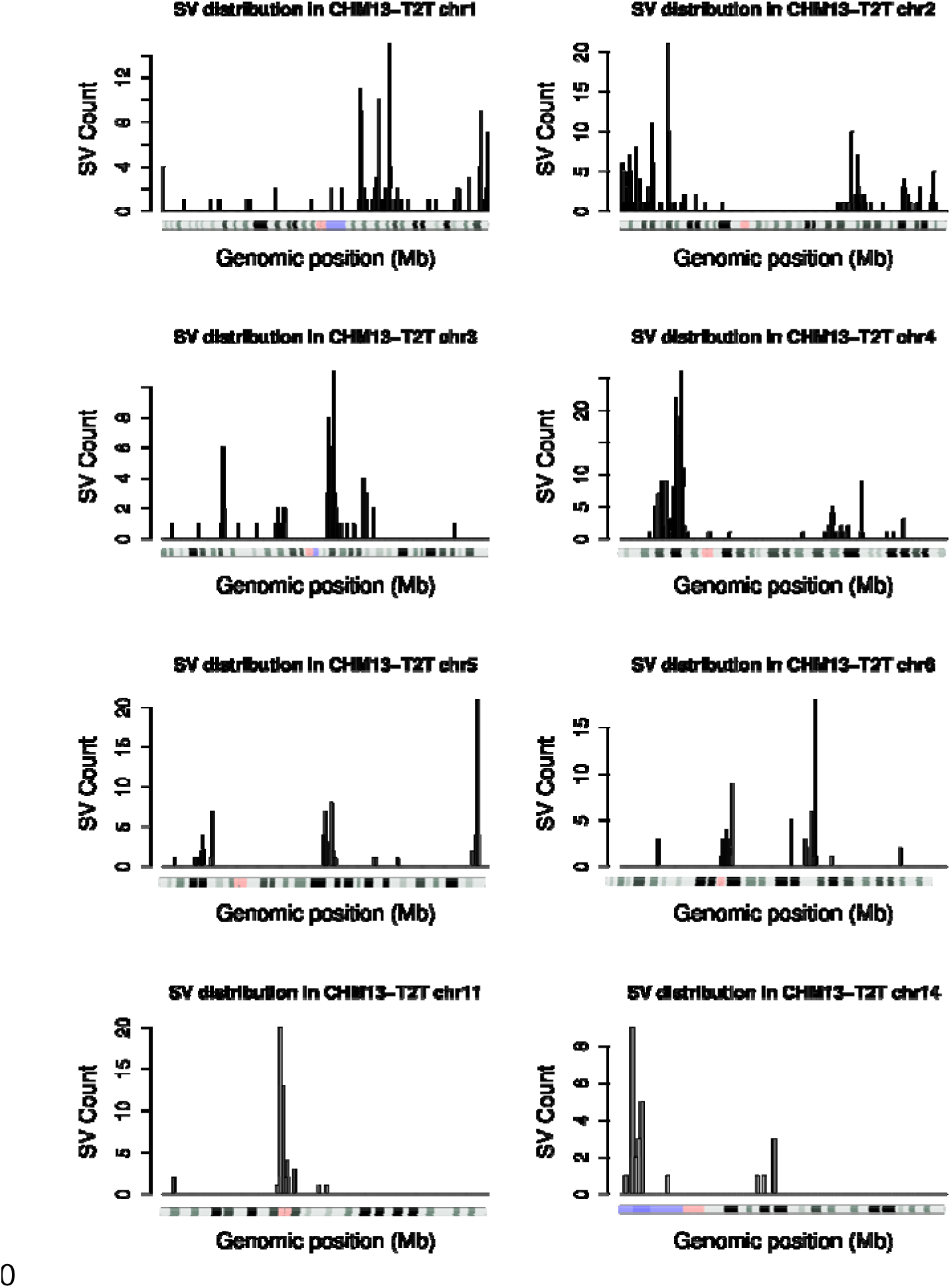
Somatic SV in cancer sample POG1022 along the CHM13-T2T genome. Shown are chromosomes 1-6, 11 and 14 where we detected a high number of somatic SVs, much of which were located in repetitive, low complexity regions.

## Discussion

In this work we assessed the benefits of using a complete reference genome for SV calling and prioritization in tumors. For this we examined four replicates of COLO829/COLO829BL together with two tumor samples aligned to both CHM13-T2T and GRCh38. We saw lower false-positive somatic SV calls when using CHM13-T2T as the reference. To enable taking advantage of CHM13-T2T mapping, while also benefiting from the richer clinical annotations available for GRCh38, we demonstrated the efficacy of lifting over alignments using LevioSAM (Chen et al. 2024). Furthermore, we developed a new approach to trace variants across reference genome versions using read names instead of coordinates, which is less sensitive to different allelic representations. Lastly, we identified inconsistencies among replicates of the existing COLO829/COLO829BL benchmark set, and propose new consensus somatic SV benchmarks for both GRCh38 and CHM13-T2T that addresses these inconsistencies. Our suggested approach should lead to more accurate and less laborious cancer SV analysis, while the improved benchmark provides a more accurate testbed for assessment of cancer SV callers.

While the advent of long reads holds promise to improve the detection of complex alleles together with resolution in tandem repeats, it also highlights the problem around variant prioritizations or annotation. In contrast to short reads, we often see many novel variants (eg. SV) that aren’t part of public databases such as gnomadSV (Collins et al. 2021), making prioritization and annotation more difficult. To overcome this, analysis often relies on tumor-normal comparison, which streamlines the detection of causative variants (Mandelker and Ceyhan-Birsoy 2020). Short reads tumour-normal analysis often indicates large (>10kbp) causative SV (Choo et al. 2023) but also has higher false positive and false negative rates (Mahmoud et al. 2019; Aganezov et al. 2022), which can hinder rapid SV prioritization during clinical workup of tumors. An example is the reporting of repeat expansions as translocation/BND events (Sedlazeck et al. 2018). We find that using long reads on different reference genomes appears to reduce the false positive calls and can thus improve variant prioritization. In this paper, we observed an important decrease in the number of BND when utilizing CHM13-T2T compared to the GRCh38 genome. Sniffles2 falsely called 130 SVs for GRCh38, 84 for CHM13-T2T and 83 for CHM13-T2T lifted over to GRCh38. False positive SV calls are often caused by misassembled and incomplete reference regions(Mahmoud et al. 2019)]; the completeness of CHM13-T2T reduces these false positives.However, clinical annotations of SVs remain incomplete for CHM13-T2T. We therefore suggest a liftover mapping approach that combines the increased accuracy of CHM13-T2T together with the utility of annotations across GRCh38 itself. Furthermore, this liftover approach does not double the analytical time nor the costs for analysis as it utilizes a chain file to rapidly liftover the read alignments without additional pairwise alignments needed(Chen et al. 2024). This also permits downstream utilization of annotation databases and approaches such as gnomadSV (Collins et al. 2021) and STIX (Chowdhury et al. 2022) to further filter and improve variant prioritization.

COLO829 and its matched control COLO829BL are well established cancer cell lines, which provide advantages for testing and benchmarking of sequencing and analysis approaches (Craig et al. 2016; Espejo Valle-Inclan et al. 2022; Pleasance et al. 2010). However, our study clearly demonstrates that caution should be used when interpreting and comparing results from a single analysis, sequencing run, or even biological sample. When we compared our benchmark to the most up to date benchmark from Valle-Inclan et al (Espejo Valle-Inclan et al. 2022), we identified multiple SVs that did not appear in any other replicates of COLO829 sequenced at other centers, and some of which were not identified in re-analysis of the original Nanopore data. For example, four SV present in the benchmark from Valle-Inclan could not be identified in any of the cancer replicates, which includes a replicate from the aforementioned benchmark. Moreover, ten SVs could not be identified in all cancer replicates or had low VAF (<10%) which complicates the interpretation of the results. This suggests caution should be used when interpreting results in comparison to the previous COLO829/COLO829BL benchmark (Espejo Valle-Inclan et al. 2022). Furthermore, across the different replicates, we could demonstrate and manually validate differences between cancer replicate samples (i.e SVs uniquely identified per replicate), similar to changes documented for somatic SNVs observed in different COLO829 passages (Craig et al. 2016). These changes clearly highlight the instability of COLO829/COLO829BL and need to be taken into consideration across benchmarks. Otherwise, there is potential for incorrect interpretation of the accuracy of sequencing and analysis techniques evaluated against the benchmark. To aid this, we identified a core set of SVs that appear to be maintained stably across independent sequencing replicates, and we propose this set to be used as the updated version of the COLO829/COLO829BL benchmark for the detection of somatic cancer SVs. It is interesting to note that some differences in the replicates of COLO829/COLO829BL can be attributed to technical artifacts, as the data set includes Nanopore data generated with prior versions of the approach that might suffer from base calling biases (ie. deletions) that have since been improved (Kolmogorov et al. 2023). Nevertheless, biological artifacts are clearly present given the evolution of the cell line.

Our work demonstrates new approaches to optimize somatic SV prioritization in cancer with potential improvements in other genetic diseases. We demonstrate this over patient samples but further over COLO829 where we introduce a new benchmark due to its variability. Given all these artifacts it’s still important to note the importance of the widely available COLO829/COLO829BL cell line for technology and analytical development.

## Methods

### Samples

We used the COLO829 cancer cell-line (melanoma) and its germline control COLO829BL (Blood, B lymphoblast) to assess somatic SV calling with whole genome long-read sequencing. We sequenced the tumor/normal samples with different technologies (ONT PromethION, ONT MinION and PacBio Revio) at different sites: Canada’s Michael Smith Genome Sciences Centre, BC, Canada (labeled GSC) sequenced with ONT PromethION; Oxford Nanopore Technologies (labeled ONT) sequenced with ONT PromethION; Pacific Biosciences of California, Inc (labeled REV) sequenced with ONT PacBio Revio; Center for Molecular Medicine and Oncode Institute, UMC Utrecht, Utrecht, the Netherlands (labeled VAI) sequenced withONT MinION.

Cancer samples were derived from the Personal Oncogenomics (POG) program (Pleasance et al. 2020), clinical trial NCT02155621, approved by and conducted under the University of British Columbia – BC Cancer Research Ethics Board (H12-00137, H14-00681). POG1022 is a tumour sample (diploid) from a metastatic diffuse large B-cell lymphoma, with the control sample taken from a skin biopsy (POG1022SK). POG044 is a tumor sample (diploid) taken from a recurrent anaplastic oligodendroglioma, with a blood control sample (POG044BL). **Supplementary Table 1** summarizes the samples used in this study and includes links for accessing the data. Two human reference genomes were utilized: GRCh38 (hg38) and CHM13-T2T (hs1).

### Alignment

Minimap2 (Li 2021) (version 2.24-r1122) was used to align the long reads to the human genome. We utilized two different versions of the human genome: GRCh38 and CHM13 version 2. We used default parameters, with the output to be in the SAM format (-a) and preset for Oxford Nanopore reads (-x map-ont). Additionally, we converted the alignment to BAM format, sorted and indexed using SAMTools (Danecek et al. 2021) (version 1.16.1).

### LiftOver

We used LevioSAM2 (Chen et al. 2024) (version 0.4.1) to liftover alignments from CHM13-T2T into GRCh38. We ran LevioSAM2 using the provided chain files for CHM13-T2Tv2. We differentiated between sequencing technologies (ONT and PacBio) in the configuration file and mapping presets for minimap2 (map-ont and ont_all.yaml for ONT and map-hifi and pacbio_all.yaml for PacBio). Additionally, we used specific allowed gaps (-g) and edit distance (-H) values for each technology: -g 1500 -H 6000 for ONT and -g 1000 -H 100 for PacBio.

### SV calling

Sniffles2 (Smolka et al. 2024) (version 2.2) was used to call structural variants (SV). Each sample had three reference backgrounds: GRCh38, CHM13-T2T, and the liftover alignment (CHM13-T2T to GRCh38). For both GRCh38 alignments (native and liftover), we added a tandem repeat annotation file during the run (--tandem-repeats file.bed). This file is provided alongside Sniffles (https://zenodo.org/records/8121996). In all cases the reference genome was provided (--reference) and the SNF file was produced (--snf file). The rest of the parameters were left as default. Next, we performed population SV calling with Sniffles2 using the SNF files produced in the previous step. Each population call was done by reference genome, such that we produced three population files, one for GRCh38, one for CHM13 and one for the liftover. In this step Sniffles2 was used with default parameters.

### Upset plot

We utilized the UpSetR packager (Conway et al. 2017) to produce upset plots that represent the number of shared SVs among samples. We used a custom script to extract the support vector provided in the VCF file (SUPP_VEC), which encodes the presence/absence of a variant in a given sample. **Supplementary tables 2, 9 and 14** contain the necessary information to produce the upset plots.

### Somatic SV detection COLO289

We used the fully-genotyped population VCF file generated in the previous step to assess the SVs that were unique to the cancer samples. We used the support vector provided in the VCF file (SUPP_VEC) to assess the presence/absence of each variant. We used bcftools (version 1.16) (Li 2011) to extract SVs that were tumor-only (using the --include option, example: bcftools view --include “SUPP_VEC = ’11110000’”). We denoted a somatic-cancer variant if the variant was only present in the cancer samples, had a VAF >= 10%, and had a minimum of 10 supporting reads. Initially, we excluded any SV that had any read support in any of the control replicates. Manual inspection overrode cases where a single read was detected in a control sample.

### SV annotation

Post-processing of SVs of the GRCh38 and CHM13-T2T to GRCh38 call sets was conducted with MAVIS (Reisle et al. 2019) (version 3.1.0). Briefly, we collapsed duplicate SVs, and merged SVs by breakpoint proximity (100 bp) and type. Next, we used the RefSeq curated gene annotation track in the UCSC genome browser (Kent et al. 2002) to annotate the impacted genes from the somatic SVs for COLO829-T2T. SV subtype-specific analyses also incorporated RepeatMasker (version 4.1) (Tarailo-Graovac and Chen 2009) to annotate the events. Events flagged as non-synonymous coding variants by MAVIS were manually reviewed.

### SV benchmark

We compared the two COLO829 somatic SV datasets that are in GRCh38 coordinates (COLO829-GRCh38 and COLO829-lifted) to a published COLO829 SV benchmark by Valle-Inclan et al. (Espejo Valle-Inclan et al. 2022), COLO829-VAI hereafter. The COLO829-VAI benchmark consists of 68 SVs, with representation from all five SV types (INS, DEL, DUP, INV, BND). We removed six SVs whose length was smaller than 50 bp (default reported by Sniffles2), leaving 62 SVs in the COLO829-VAI benchmark. We used BEDtools (version 2.31) (Quinlan and Hall 2010) to compare the SV coordinates of the COLO829-VAI benchmark to our COLO829-GRCh38 and COLO829-lifted somatic SV datasets.

For the case of the COLO829-T2T dataset, we included the --output-rnames parameter in Sniffles to output the read names supporting each SV. We then used these read names along with the chromosome and the SV type as a proxy for matching the SV from CHM13-T2T to GRCh38 coordinates to perform a partial benchmark (FP only). Only for one case we used the same procedure to investigate a FN call in the COLO829-T2T dataset (DEL in chr16), because Sniffles made the call and reported the read names. However, the SV was removed from the final call set by the COV_MIN filter, thus it was still considered a FN.

### SV genotyping (force-calling)

We used the Sniffles2 --genotype-vcf option to look for all the FN calls in the COLO829-GRCh38 and COLO829-lifted datasets. This option takes a VCF as input (including the FN calls for our case) and uniquely searches for the SV present in the VCF input and updates the genotype according to what Sniffles detects in the alignment file. For the case of BNDs, we additionally took all the reads overlapping with the coordinates provided by the COLO829-VAI benchmark and searched for reads that had supplementary alignments (SA; flag 2048) and compared the coordinates from the SA tag in the alignment to the “CHR2” value from the INFO field of the COLO829-VAI benchmark. We matched chromosomes and allowed for a 10kb distance between the positions.

### Somatic SV detection POG samples

We used the fully-genotyped population VCF file generated by merging tumor and normal samples using Sniffles2 SNF files (sniffles --input tumor.snf normal.snf --vcf merge.vcf.gz). Once merged, we used the support vector provided in the VCF file (SUPP_VEC) to assess the presence/absence of each SV. We used bcftools (version 1.16) to extract SVs that were tumor-only (using the --include option, example: bcftools view --include “SUPP_VEC = ’10’”). We denoted a somatic-cancer variant if the SV was only present in the cancer samples, a VAF >= 10% and a minimum of 10 read support.

### Data access

**Supplementary Table 1** summarizes the samples used in this study and includes links for accessing the data. The new COLO829/COLO829BL proposed benchmark and merge VCF files that include the eight tumor/normal samples can be found at 10.5281/zenodo.10819636. Nutty, a Sniffles2 companion app for parsing the VCF was used https://github.com/lfpaulin/nutty It contains commands to reproduce the COLO829 and POG analysis.

## Data Availability

Supplementary Table 1 summarizes the samples used in this study and includes links for accessing the data. The new COLO829/COLO829BL proposed benchmark and merge VCF files that include the eight tumor/normal samples can be found at 10.5281/zenodo.10819636. Nutty, a Sniffles2 companion app for parsing the VCF was used https://github.com/lfpaulin/nutty It contains commands to reproduce the COLO829 and POG analysis.

## Acknowledgements

This study was in part supported by funding from the Canada Research Chairs Program, Terry Fox Research Institute Marathon of Hope and the British Columbia Cancer Foundation. FJS, LFP was supported by NIH (UM1DA058229, 1UG3NS132105-01, 1U01HG011758-01). This study was conducted with the financial support of The Terry Fox Research Institute and the Terry Fox Foundation. The views expressed in the publication are the views of the authors and do not necessarily reflect those of the Terry Fox Research Institute or the Terry Fox Foundation.

